# From Screening to Sustained Recovery: A Multidomain Systematic Review and Evidence Map of Adolescent Substance-Use Rehabilitation with Nested Meta-analysis of Youth Opioid Treatment

**DOI:** 10.64898/2026.07.11.26357831

**Authors:** Prateek Mittal, Ayush Srivastava, Prabal Pratap Singh, Joohi Chauhan

**Affiliations:** Vision Exploration and Data Analytics (VEDAs) Lab, Motilal Nehru National Institute of Technology Allahabad, India; Vellore Institute of Technology, Vellore, India; Foundation for Ethical AI in Healthcare (FEAIH), Faridabad, India

## Abstract

**Background:** Adolescent substance-use rehabilitation is a care-continuum problem spanning detection, engagement, active treatment, relapse prevention, aftercare, family support, and equity-oriented implementation. Existing reviews are often modality-specific and do not show how evidence aligns with substances, populations, outcomes, stages of care, or policy needs.

**Objectives:** To map and synthesise the 2015-2025 adolescent and transitional-age youth SUD rehabilitation literature across intervention domains, stages, substances, outcomes, equity/disadvantage, geography, and economics, and to perform meta-analysis only where pooling was clinically defensible.

**Methods:** PubMed, Scopus, and Web of Science records were harmonised to 2015-2025 and deduplicated. Two reviewer roles applied a predefined charting codebook for substance focus, technique family, rehabilitation stage, equity/disadvantage flags, outcome family, and study-design signal. Evidence was synthesised across AI/digital, psychiatric/psychotherapeutic, pharmacological, family/social, behavioural, residential/continuing-care, school/community, harm-reduction, and policy domains. Random-effects meta-analysis was restricted to comparative youth OUD medication-supported trials with extractable binary outcomes.

**Results:** The search identified 1,676 records; 554 duplicates were removed, leaving 1,122 unique records. Metadata screening retained 579 records for evidence-map charting: 112 high-confidence records and 467 conservative metadata-supported records requiring full-text verification before final selective-journal submission. The charted evidence was concentrated in active treatment (n=433) and relapse prevention (n=114); aftercare/follow-up was weak (n=8). Intervention-family signals were led by pharmacological/MOUD (n=72), psychotherapy/psychiatric care (n=65), school/community/brief interventions (n=46), residential/continuing care (n=41), family/social therapy (n=30), AI/digital/telehealth (n=25), harm-reduction/policy (n=24), and CM (n=22). The primary youth OUD retention/completion meta-analysis favoured medication-supported treatment (OR 7.67, 95% CI 3.98-14.78; I^2^=0%; k=2; n=188). An exploratory favourable-outcome analysis produced a similar estimate (OR 7.94, 95% CI 4.24-14.89; I^2^=0%; k=3; n=229).

**Conclusions:** The strongest pooled quantitative claim supports medication-supported treatment for youth OUD. For non-opioid substances, digital care, family therapy, CM, residential care, aftercare, and equity-oriented implementation, the literature is clinically important but not yet consistently synthesis-ready. Future trials should evaluate complete care pathways, adopt core outcomes, report age-banded and equity subgroup effects, and include economic and implementation endpoints.

**Research in context:** *Evidence before this study:* Existing adolescent SUD reviews are often organised by single intervention families: outpatient behavioural treatment, family therapy, brief interventions, alcohol-focused psychosocial treatment, pharmacological treatment for youth OUD, or digital interventions[1, 2, 3, 4, 5, 6, 7, 8, 9, 10, 11, 12]. That work is valuable, but it does not fully answer rehabilitation-system questions: where young people are detected, how they are engaged, which treatment layer is matched to which substance, what happens after discharge, and whether evidence is equitable across gender, ethnicity, justice involvement, geography, and cost.

*Added value of this study:* We analyse adolescent SUD rehabilitation as a staged care-continuum and multidomain evidence system. The synthesis combines a harmonised search, PRISMA 2020-style flow, dual-reviewer charting supplement, technique-by-stage matrices, substance-by-technique matrices, outcome-gap matrices, geographic mapping, and nested meta-analysis for the only subgroup in which pooling is clinically defensible: medication-supported youth OUD treatment.

*Implications:* The clearest pooled quantitative signal supports medication-supported treatment for youth OUD. For alcohol, cannabis, stimulants, digital tools, family therapy, residential care, aftercare, and equity-oriented implementation, the major problem is not absence of ideas; it is fragmented outcomes, underpowered subgroup analyses, weak aftercare measurement, and limited economic and global-south evidence.

## 1 Introduction

Adolescent and transitional-age youth SUDs are not reducible to a single drug, programme, or end-point. They sit at the intersection of neurodevelopment, school functioning, family conflict, psychiatric comorbidity, peer networks, stigma, poverty, rurality, juvenile justice, and rapidly changing drug markets[13, 14, 15, 16, 17]. A young person may enter care through paediatrics, psychiatry, school counselling, family referral, emergency care, outreach, or the justice system. A rehabilitation review must therefore ask not only whether an intervention reduces use during an episode, but also whether it improves detection, engagement, retention, relapse prevention, aftercare, family functioning, education or work reintegration, and equity of access.

The evidence base is unusually multidomain. Psychiatric and psychotherapeutic approaches include MI, MET, CBT, brief behavioural interventions, and comorbidity-focused care[2, 3, 6, 7, 18]. Pharmacological evidence is strongest for OUD, where buprenorphine, naltrexone, and MOUD delivery models are central, but youth access remains limited by stigma, licensing, family consent, service availability, and treatment discontinuity[8, 9, 10, 19, 20]. Family and social therapies, including MDFT, FFT, MST, CRAFT, and parent-training components, are developmentally aligned but difficult to scale[4, 5, 21, 22]. CM is mechanistically strong but constrained by funding and implementation rules[23, 24]. Digital and AI tools are expanding, but the adolescent SUD literature is still early and safety concerns are substantial[25, 11, 12, 26, 27, 28, 29].

This breadth explains why ordinary scoping reviews of adolescent rehabilitation are vulnerable to desk rejection: they list modalities but do not produce an integrative analytic story. Conversely, a narrow meta-analysis misses the pathway-level and equity questions that determine whether evidence can be implemented. We therefore designed this review as a systematic evidence map with multidomain synthesis and nested meta-analysis only where pooling was clinically defensible.

### 1.1 Objectives

The review had five objectives: (1) map the 2015-2025 adolescent and transitional-age youth SUD rehabilitation literature across substances, intervention families, care stages, outcomes, and equity domains; (2) evaluate whether the literature is concentrated in particular stages of rehabilitation; (3) examine substance-intervention mismatch, especially whether pharmacological evidence generalises beyond youth OUD; (4) identify gaps in gender, race/ethnicity, justice involvement, socioeconomic disadvantage, rurality, LMIC/global-south representation, and economic evaluation; and (5) statistically re-evaluate effect-level claims only where a clinically coherent subgroup allowed defensible pooling.

### 1.2 Paper organisation

After Methods, the Results proceed as a story rather than a catalogue: first, search yield and charting profile; second, care-continuum architecture; third, technique-stage and substance-technique matrices; fourth, outcome and equity gaps; fifth, domain-specific synthesis; sixth, nested meta-analysis. The Discussion then converts these findings into implementation priorities, a core outcome set, and a research agenda.

## 2 Methods

### 2.1 Design and reporting standards

We used a hybrid design: a systematic evidence map to describe the multidomain rehabilitation literature and a nested meta-analysis restricted to youth OUD medication-supported interventions. Reporting was guided by PRISMA 2020 for the systematic review component and PRISMA-ScR for the evidence-map logic[30, 31]. Because this study maps heterogeneous intervention families while pooling only one coherent clinical subgroup, we explicitly separate map-level claims from effect-level claims.

### 2.2 Search sources and dates

PubMed, Scopus, and Web of Science were searched for records from 1 January 2015 to 31 December 2025. The Scopus and Web of Science search strings supplied for this project used a 2015-2025 publication-year window; PubMed records beyond 2025 were therefore removed before analysis to maintain cross-database consistency. Search strings combined terms for adolescents/youth, SUD or specific substances, rehabilitation/ treatment/ intervention modalities, and outcomes. Exact search strings are provided in Appendix A.

### 2.3 Eligibility framework

### 2.4 Dual-reviewer charting and extraction

Two reviewer roles, Prateek and Ayush, independently screened and charted records using a predefined codebook. Reviewer 1 prioritised sensitivity for adolescent rehabilitation relevance; Reviewer 2 prioritised specificity for direct SUD rehabilitation relevance. For the revised supplement, reviewer decisions were simplified to chart/do-not-chart votes, and all retained records were labelled by evidence-map confidence and full-text-verification requirement rather than by ambiguous interim categories. The supplementary workbook provides record-level provenance, charting votes, extraction fields, charting codes, and verification flags.

### 2.5 Synthesis domains

Extraction was organised around five axes: technique family, care stage, substance focus, outcome family, and equity/disadvantage. To keep multi-panel figures readable, short codes were used in heatmaps and matrices; their expansions are provided in Table 3. Categories were non-mutually exclusive because adolescent rehabilitation programmes often combine modalities.

**Table 2.**
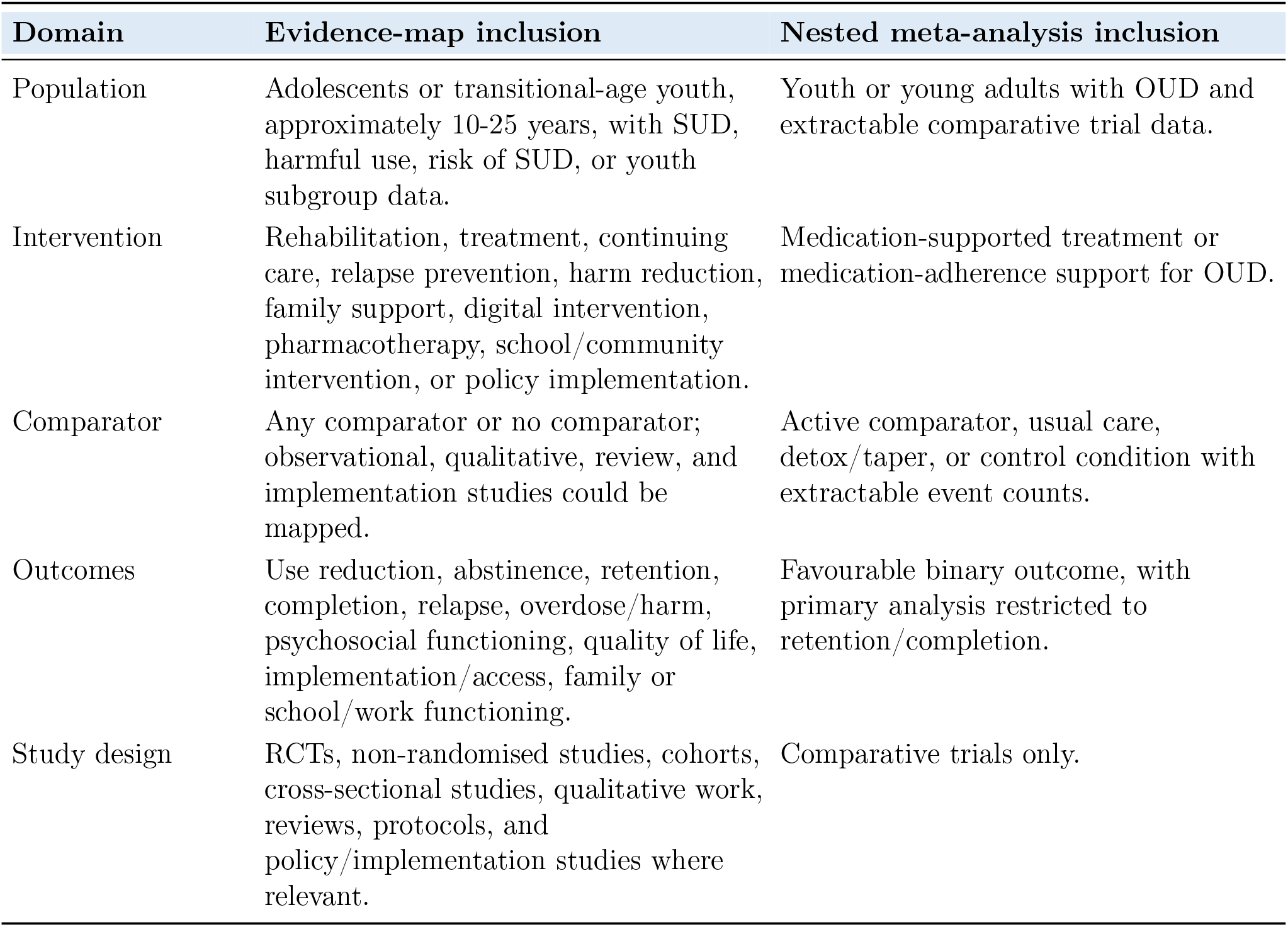
Eligibility framework for the evidence map and nested meta-analysis.

| Domain | Evidence-map inclusion | Nested meta-analysis inclusion |
| --- | --- | --- |
| Population | Adolescents or transitional-age youth, approximately 10-25 years, with SUD, harmful use, risk of SUD, or youth subgroup data. | Youth or young adults with OUD and extractable comparative trial data. |
| Intervention | Rehabilitation, treatment, continuing care, relapse prevention, harm reduction, family support, digital intervention, pharmacotherapy, school/community intervention, or policy implementation. | Medication-supported treatment or medication-adherence support for OUD. |
| Comparator | Any comparator or no comparator; observational, qualitative, review, and implementation studies could be mapped. | Active comparator, usual care, detox/taper, or control condition with extractable event counts. |
| Outcomes | Use reduction, abstinence, retention, completion, relapse, overdose/harm, psychosocial functioning, quality of life, implementation/access, family or school/work functioning. | Favourable binary outcome, with primary analysis restricted to retention/completion. |
| Study design | RCTs, non-randomised studies, cohorts, cross-sectional studies, qualitative work, reviews, protocols, and policy/implementation studies where relevant. | Comparative trials only. |

**Table 3.** Code legend used in synthesis figures.

| Dimension | Codes |
| --- | --- |
| Technique | DIG=AI/digital/telehealth; PSY=Psychotherapy/psychiatric; PHA=Pharmacological/MOUD; FAM=Family/social therapy; CM=Behavioural incentives/contingency management; RES=Residential/continuing care; SCH=School/community/brief; HRP=Harm reduction/policy. |
| Care stage | DET=Discovery/detection; ENG=Engagement/initiation; TX=Active treatment; REL=Relapse prevention; AFT=Aftercare/follow-up; FAM-S=Family support/counselling; EQU=Policy/equity implementation. |
| Substance | OPI=Opioids; ALC=Alcohol; CAN=Cannabis; STI=Stimulants; NIC=Tobacco/nicotine; POLY=General/polysubstance. |
| Outcome | USE=Abstinence/use reduction; RET=Retention/completion; HARM=Relapse/overdose/harm; FUNC=Psychiatric/functioning; ACCESS=Implementation/access. |

**Table 4.**
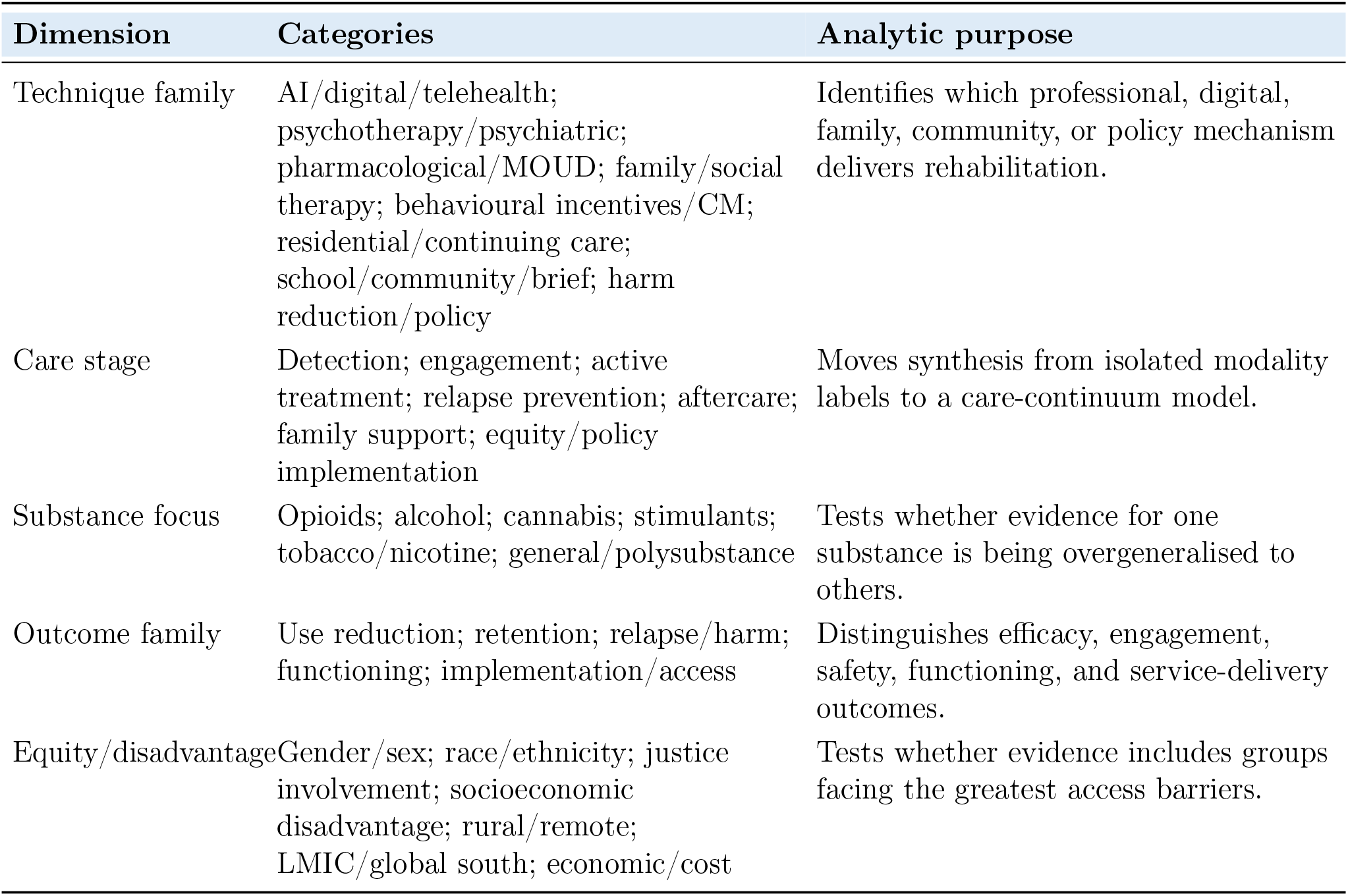
Multidomain extraction taxonomy.

**Table 5.**
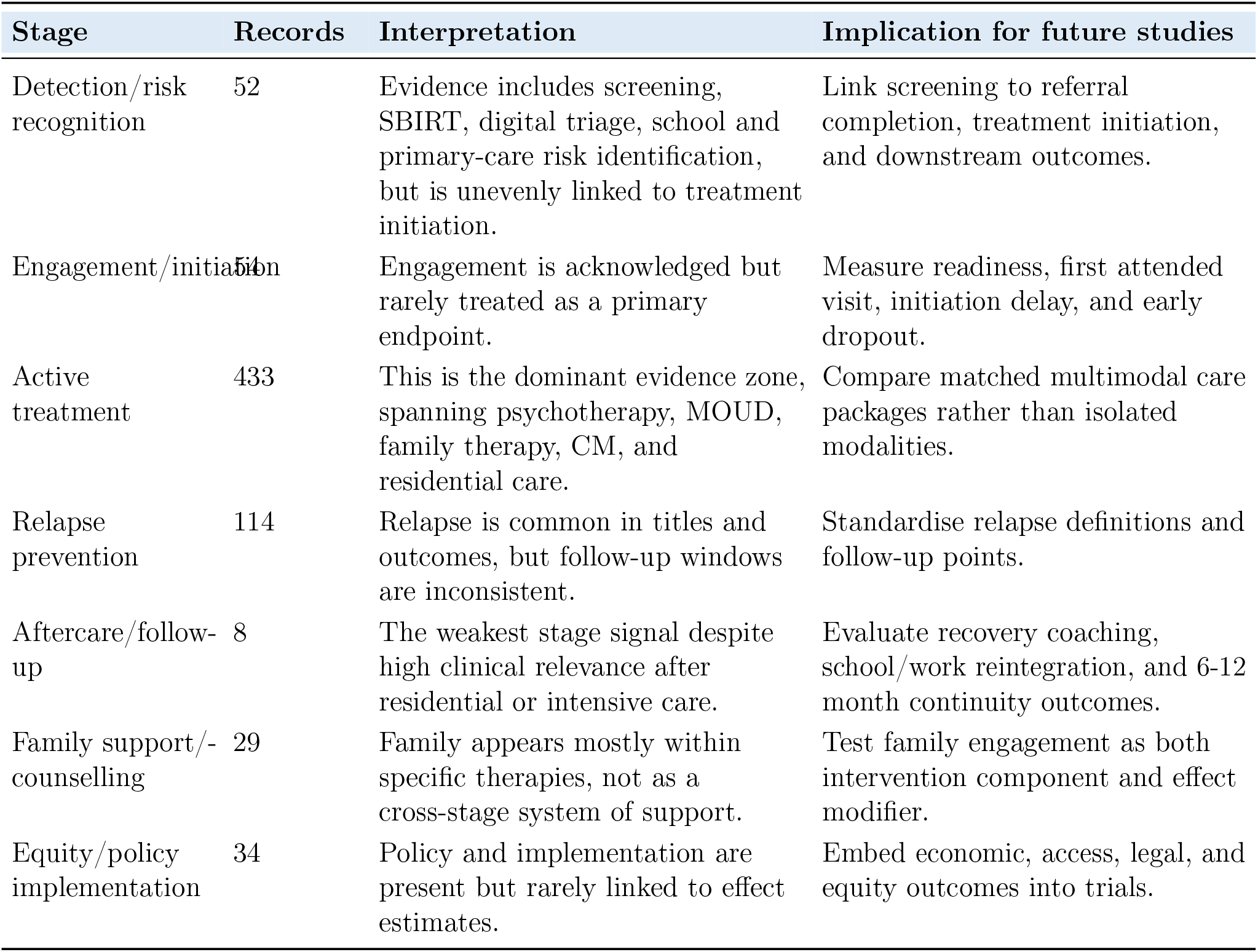
Stage-specific interpretation of the evidence map.

**Table 6.** Technique-specific synthesis across adolescent SUD rehabilitation.

| Domain | Records | Current status | Main limitation | Actionable interpretation |
| --- | --- | --- | --- | --- |
| AI/digital/telehealth | 25 | Apps, web programmes, SMS, telehealth, relapse prediction, and AI-assisted tools are expanding[25, 11, 12, 26, 27, 35, 28, 36, 37, 38]. | Short follow-up, feasibility focus, crisis-safety uncertainty, algorithmic bias, and device/access inequity. | Use as adjunctive engagement and monitoring tools until adolescent-specific effectiveness and safety trials mature. |
| Psychotherapy/psychiatric | 65 | CBT, MI/MET, brief interventions, and comorbidity care remain central for non-opioid SUD and outpatient treatment[2, 3, 6, 7, 18, 13, 39, 40, 41]. | Heterogeneous comparators, dose, fidelity, therapist training, and outcome windows. | Treat as core psychosocial layer; require harmonised outcomes and comorbidity stratification. |
| Pharmacological/MOUD | 72 | Strongest quantitative evidence is for OUD, especially buprenorphine and medication-supported care[8, 42, 10, 43, 9, 44, 45, 46, 47, 48, 49, 19, 20]. | Evidence does not generalise to cannabis, alcohol, stimulants, or polysubstance care. | Implement youth OUD MOUD as standard care; do not infer equivalent pharmacological maturity for non-opioid SUD. |
| Family/social therapy | 30 | MDFT, FFT, MST, parent training, and community reinforcement models align well with adolescent developmental context[4, 5, 21, 22, 50, 51, 52, 53]. | Resource intensity, workforce needs, cultural adaptation, and family availability. | Scale with fidelity monitoring and cultural tailoring; report family functioning and caregiver engagement. |
| Behavioural incentives/CM | 22 | CM has a strong reinforcement mechanism and trial support, especially for cannabis and adherence contexts[23, 24, 34]. | Reimbursement barriers, ethical concerns, and implementation variability. | Treat as high-value but policy-constrained; evaluate pragmatic CM in community and family-supported models. |
| Residential/continuing care | 41 | Residential care, continuing care, recovery coaching, and reintegration appear across the literature[54, 55, 56, 57, 58, 59, 60]. | Admission and completion often measured more than post-discharge recovery. | Require aftercare linkage, relapse, school/work, housing, and 6-12 month outcomes. |
| School/community brief | 46 | School, primary-care, SBIRT, outreach, and brief intervention models offer reach[61, 62, 63, 6, 16]. | Referral completion and treatment linkage are often weakly measured. | Evaluate the full pathway from detection to first treatment contact and retention. |
| Harm reduction/policy | 24 | Naloxone, justice diversion, anti-stigma work, decriminalisation, and access policy are critical for equity[64, 16, 65, 66, 67, 17]. | Youth-specific evidence, legal contexts, and acceptability vary widely. | Frame harm reduction as a bridge to care, not a competing philosophy. |

**Table 7.** Substance-specific interpretation and synthesis needs.

| Substance focus | Dominant evidence pattern | What can be claimed now | What remains weak |
| --- | --- | --- | --- |
| Opioids (n=116) | Medication-supported care, retention, overdose/harm reduction, and stigma/access barriers. | Youth OUD should not be treated with psychosocial care alone when MOUD is appropriate. | Youth-specific initiation, retention, family consent, and rural access models. |
| Alcohol (n=95) | Brief interventions, CBT/MI/MET, school/community and psychosocial interventions. | Psychosocial and brief interventions are the clearest first-line evidence families. | Long-term relapse and functional recovery outcomes. |
| Cannabis (n=63) | CBT/MI/MET, CM, family therapy; weak pharmacotherapy signal. | Behavioural and family/social models are more actionable than medication claims. | Standardised cannabis outcomes and comorbidity stratification. |
| Stimulants (n=9) | Behavioural, community, and harm-reduction approaches. | No strong adolescent pharmacotherapy claim can be made from this corpus. | Trials of CM, psychiatric comorbidity care, and harm reduction. |
| Nicotine/tobacco (n=36) | Behavioural plus pharmacological cessation themes. | Combination models may be relevant but require youth-specific adherence data. | Vaping-specific recovery, relapse, and dual-use outcomes. |
| Poly/general (n=266) | Mixed modality, treatment-system, and implementation literature. | Multimodal assessment and matched care are necessary. | Substance-specific effect estimates and subgroup reporting. |

**Table 8.** Proposed minimum core outcome set for adolescent SUD rehabilitation studies.

| Outcome domain | Minimum reporting item | Why it matters |
| --- | --- | --- |
| Engagement | Referral completion; first attended visit; time to initiation. | Screening without initiation does not constitute rehabilitation. |
| Retention | Days retained; visits attended; programme completion; medication continuation. | Retention is a major determinant of youth outcomes and meta-analytic comparability. |
| Use reduction | Substance-specific frequency; abstinence; biologically verified outcomes where feasible. | Different substances require distinct definitions. |
| Relapse and harm | Relapse; overdose; emergency visits; justice contact; unsafe-use episodes. | Particularly important in fentanyl and polysubstance contexts. |
| Mental health and functioning | Depression/anxiety symptoms; family functioning; school/work participation; quality of life. | Recovery must be developmental and functional, not only toxicological. |
| Aftercare | Continuing-care linkage; recovery coaching; family follow-up; school/work reintegration. | The weakest part of the current evidence map. |
| Equity and cost | Age band; sex/gender; race/ethnicity; justice involvement; rurality; socioeconomic status; cost. | Required to determine who benefits and who remains excluded. |

**Table 9.**
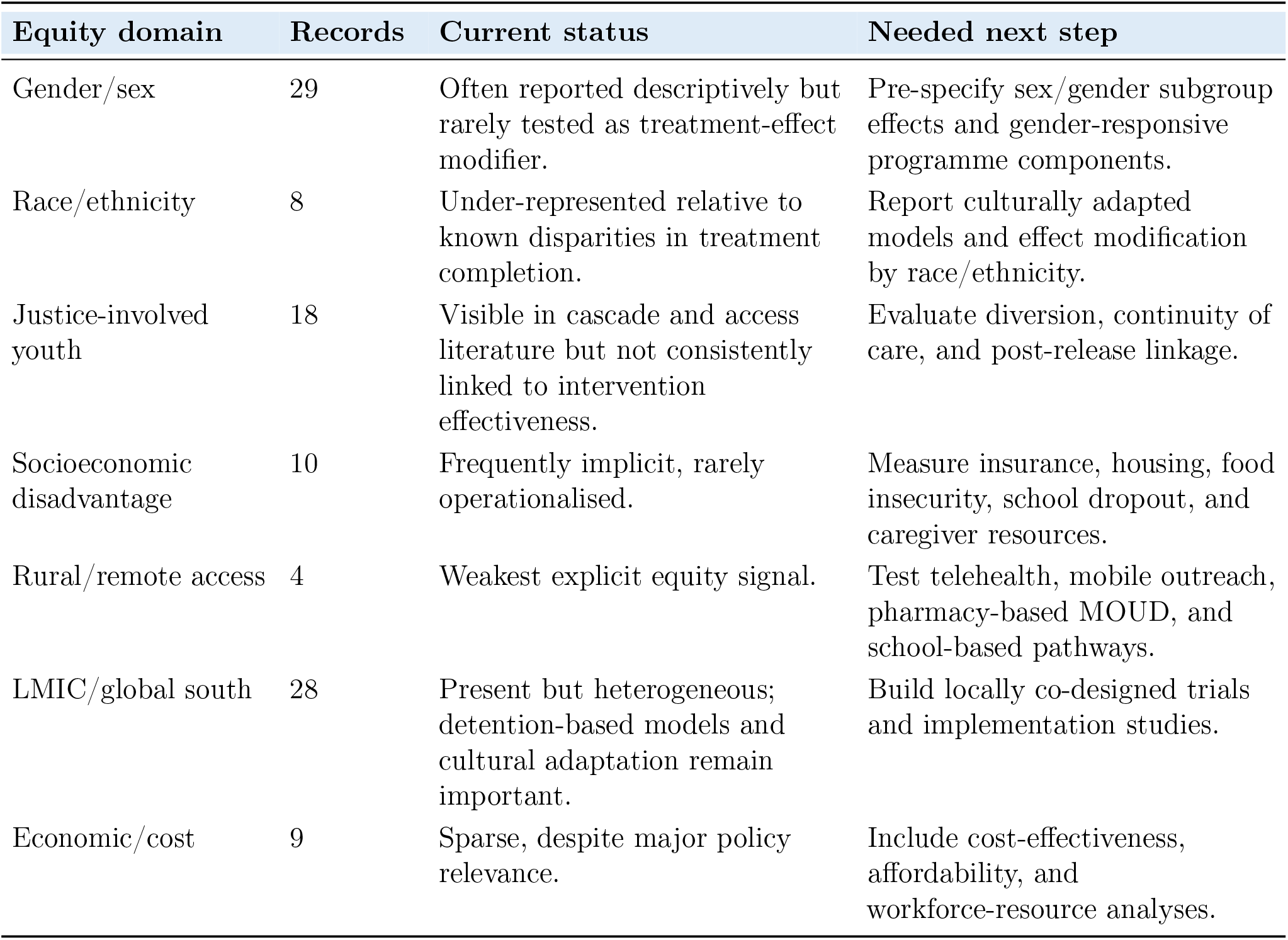
Equity and implementation synthesis.

**Table 10.** Trials contributing to the nested youth OUD meta-analysis.

| Study | Intervention | Comparator | Outcome used | Intervention | Control |
| --- | --- | --- | --- | --- | --- |
| Marsch et al. 2005[32] | Buprenorphine plus counselling | Clonidine plus counselling | Completed 28-day detoxification | 13/18 | 7/18 |
| Woody et al. 2008[33] | 12-week buprenorphine-naloxone | 14-day taper/detoxification | Retained at week 12 | 52/74 | 16/78 |
| Fishman et al. 2021[34] | Youth opioid recovery support plus XR-NTX adherence support | Treatment as usual | No relapse at week 24 | 8/21 | 1/20 |

**Table 11.** Statistical claim audit: what this review can and cannot infer.

| Claim type | Supported interpretation | Boundary condition |
| --- | --- | --- |
| Medication-supported youth OUD treatment improves retention/completion | Supported by nested pooled estimate favouring medication-supported care. | OUD-specific; small number of youth trials; not a general adolescent SUD pharmacotherapy claim. |
| Psychotherapy is central for non-opioid SUD | Supported by large and updated behavioural-treatment literature. | Outcome heterogeneity limits a single pooled effect across alcohol, cannabis, stimulant, nicotine, and polysubstance presentations. |
| Digital/AI tools can extend reach | Plausible and increasingly studied. | Clinical efficacy, crisis safety, bias, and adolescent-specific regulatory readiness remain under-validated. |
| Family therapy is developmentally aligned | Supported by family-therapy reviews and selected trials. | Scaling, workforce, family availability, and cultural adaptation are major constraints. |
| Aftercare is essential | Strong clinical rationale and some continuing-care evidence. | Severely under-measured in the charted corpus. |
| Equity is a structural gap | Supported by extraction signals and disparity literature. | Many records report demographics descriptively rather than testing subgroup effects. |

**Table 12.** Policy and research implications from the multidomain synthesis.

| Area | Current status | Recommended action |
| --- | --- | --- |
| Youth OUD | Medication-supported care has the strongest pooled signal but remains underused. | Remove access barriers, support paediatric/adolescent prescribers, integrate family and psychosocial care, and track retention. |
| Non-opioid SUD | Psychosocial and family/community interventions dominate; medication evidence is weaker. | Harmonise outcomes and test combined behavioural-family-CM packages by substance. |
| Digital/AI | Growing but not yet mature for stand-alone care. | Require adolescent-specific trials, human oversight, crisis escalation, bias audits, and privacy safeguards. |
| Aftercare | Severely underrepresented despite relapse risk after discharge. | Fund continuing-care trials with 6- and 12-month outcomes and school/work reintegration metrics. |
| Equity | Often descriptive rather than analytic. | Pre-specify subgroup analyses and co-design interventions with justice-involved, rural, LMIC, and marginalised youth. |
| Economics | Sparse despite policy importance. | Add cost-effectiveness, affordability, workforce, and reimbursement outcomes to pragmatic trials. |
| Reporting | Outcome heterogeneity prevent synthesis. | Adopt the core outcome set proposed in Table 8. |

**Table 13.**
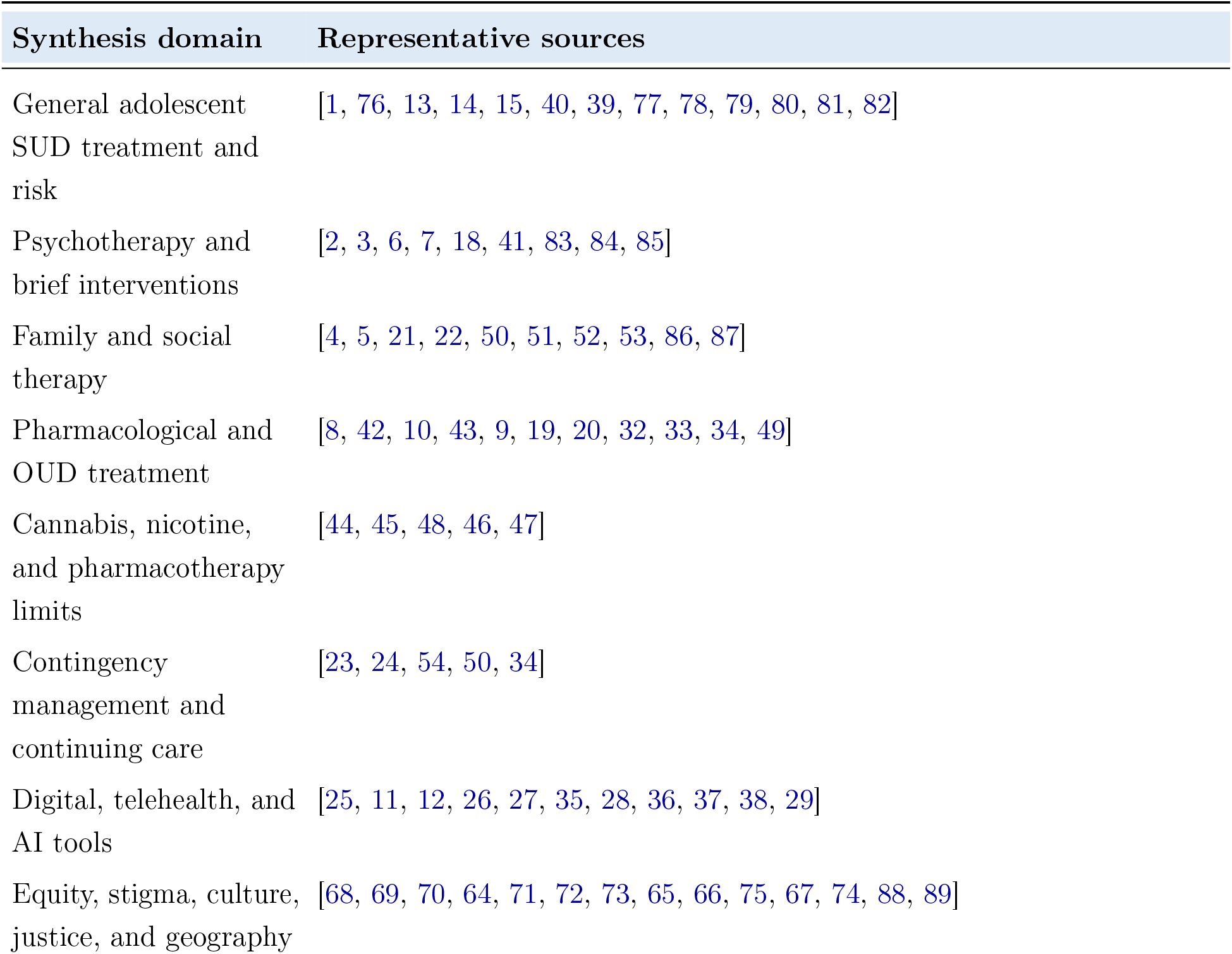
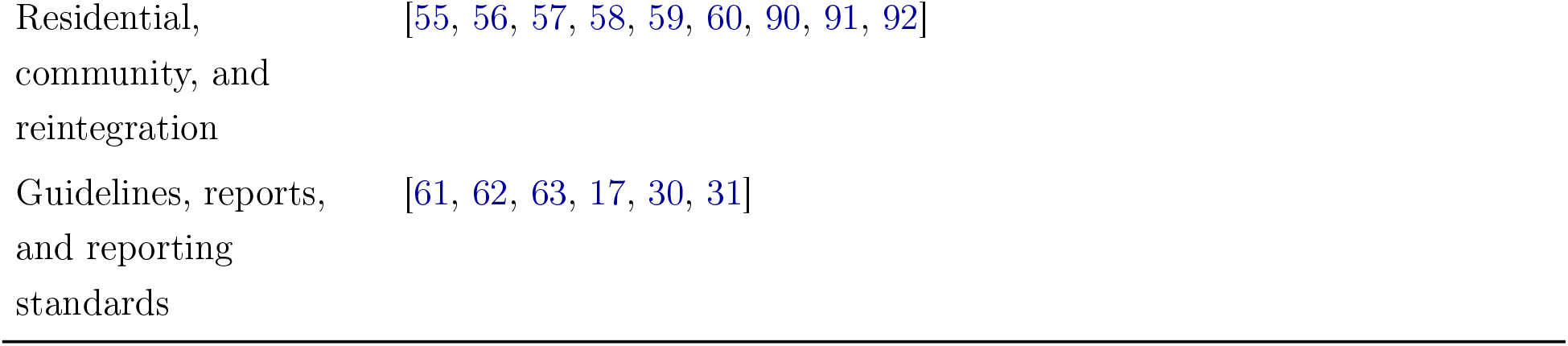
Representative literature cited to support the multidomain synthesis. This table is not intended to list all 579 charted records; the full record-level supplement provides that audit trail.

### 2.6 Statistical analysis

For the nested OUD meta-analysis, event counts were extracted from comparative youth OUD medication supported trials. ORs were calculated such that values greater than 1 favoured medication-supported intervention. The primary analysis pooled treatment completion/retention outcomes from Marsch et al. and Woody et al.[32, 33]. An exploratory analysis added Fishman et al.[34], which tested youth opioid recovery support and medication-adherence support with a relapse-related favourable outcome. Random-effects inverse-variance pooling was used; heterogeneity was summarised with *I*^2^ and *τ* ^2^. Small-study-effect tests were not performed because fewer than ten studies were pooled.

## 3 Results

### 3.1 Search yield and PRISMA flow

The harmonised database search identified 1,676 records: PubMed (n=722), Scopus (n=685), and Web of Science (n=269). Deduplication removed 554 records, leaving 1,122 unique records for screening. Metadata screening excluded 543 records and retained 579 for evidence charting. Within this charted set, 112 records had sufficient metadata specificity for high-confidence charting, and 467 were conservatively retained as metadata-supported records requiring full-text verification. This structure is important: a mature top-journal submission should not hide uncertainty caused by missing abstracts or mixed-age samples. Instead, it should expose the verification requirement without using ambiguous screening labels.

### 3.2 Care-continuum architecture

Adolescent rehabilitation evidence is heavily concentrated in the middle of the pathway: active treatment. In the charted corpus, active treatment appeared in 433 records, relapse prevention in 114, engagement/initiation in 54, and detection in 52. By contrast, aftercare/follow-up appeared in only 8 records and family support/counselling in 29. This is not a minor imbalance. Adolescents often do not self-refer, often disengage early, and often relapse after discharge. Therefore, the stages least represented in the literature are precisely the stages most likely to determine whether rehabilitation becomes durable.

### 3.3 Technique-family synthesis

The most frequent intervention-family signals were pharmacological/MOUD (n=72), psychotherapy/psychiatric care (n=65), school/community/brief interventions (n=46), residential/continuing care (n=41), family/social therapy (n=30), AI/digital/telehealth (n=25), harm-reduction/ policy (n=24), and behavioural incentives/CM (n=22). However, volume did not equal maturity. Psychotherapy and MOUD were more trial-visible, while digital, policy, after-care, and equity work were more often feasibility, implementation, or observationally oriented.

### 3.4 Technique by stage matrix

Figure 4 shows that active treatment dominates most intervention families. Detection is mainly tied to school/community/brief and digital approaches. Aftercare is weak across nearly all technique families. Family support appears both as a named therapy family and as a care stage, but its cross-stage role is underdeveloped. This suggests that adolescent rehabilitation research has not yet fully adopted a longitudinal recovery model.

**Figure 1.**
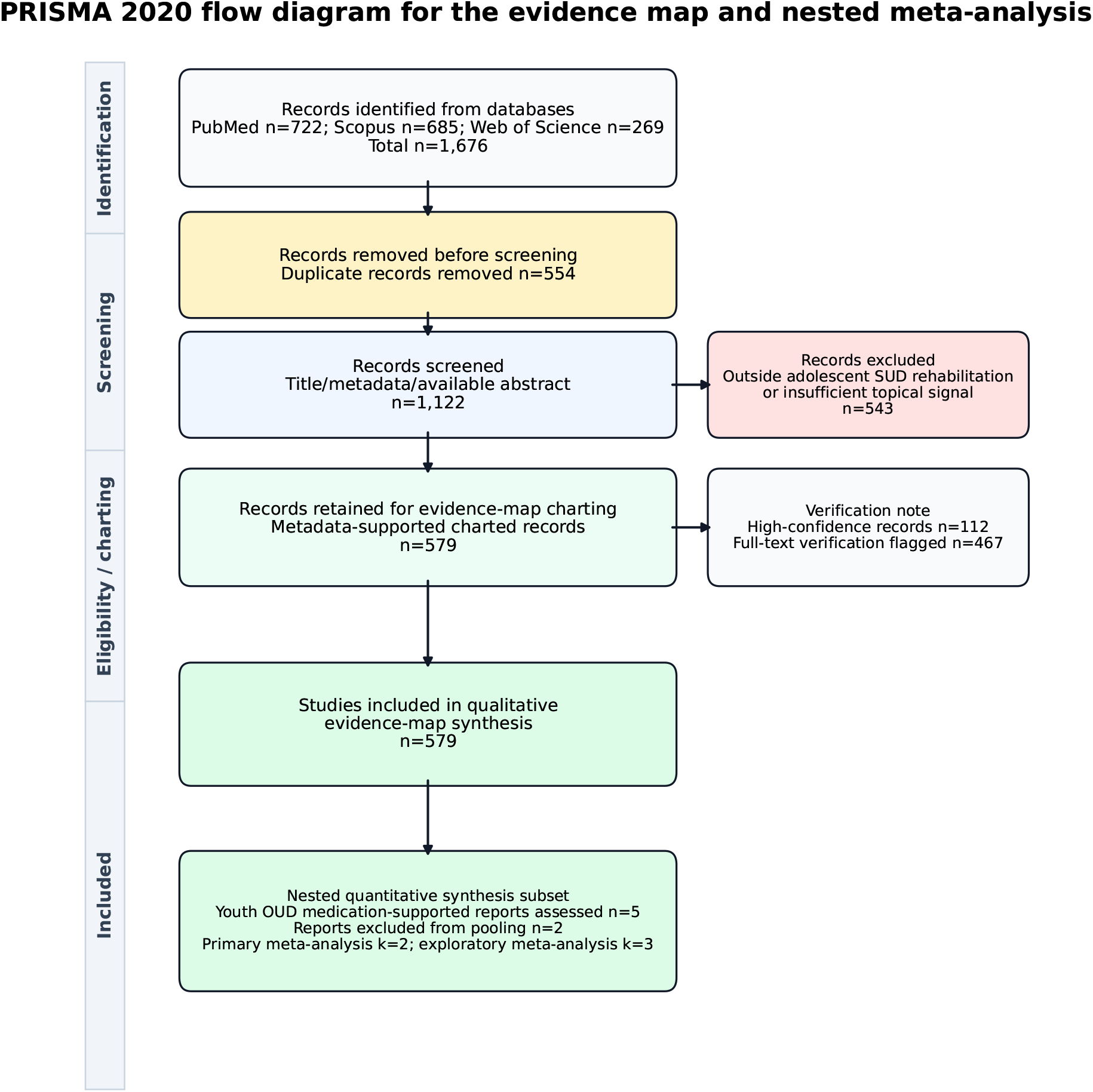
PRISMA 2020 flow diagram for the evidence-map and nested meta-analysis components. The qualitative evidence map retained 579 metadata-supported records, while the nested quantitative synthesis was restricted to comparative youth OUD medication-supported trials with extractable binary outcomes.

**Figure 2.**
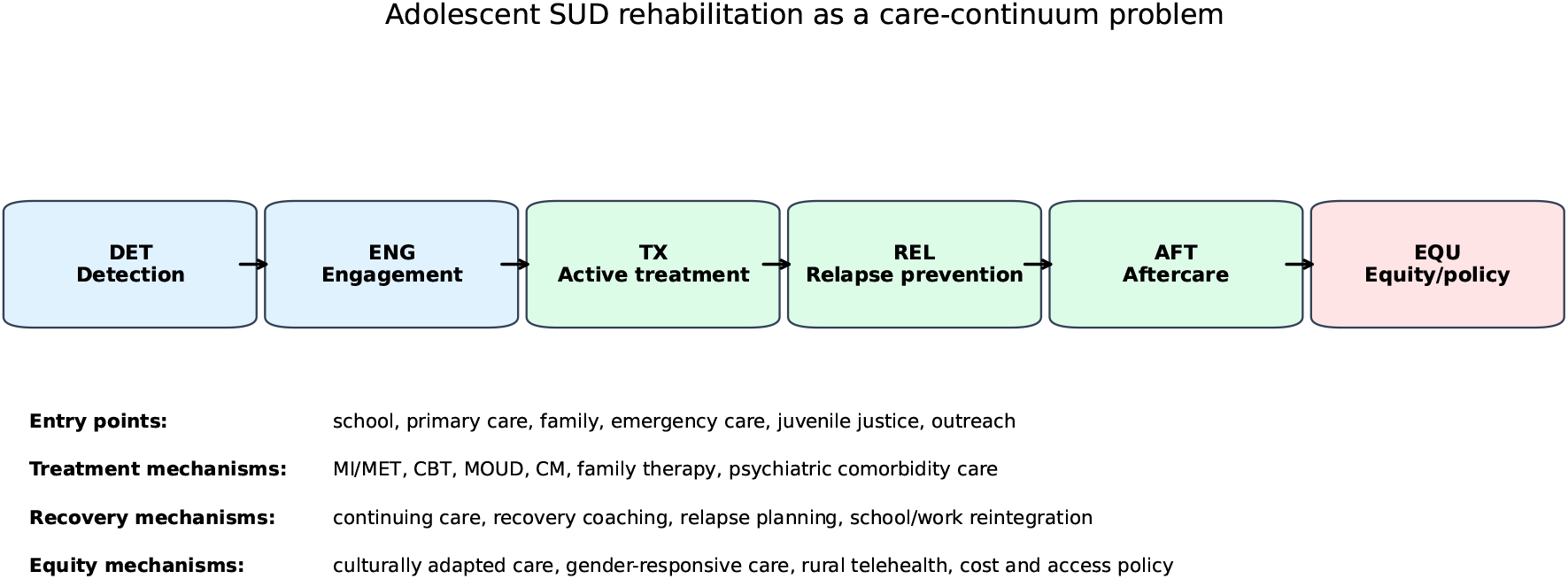
Care-continuum framing for adolescent SUD rehabilitation. The review separates evidence for detection, engagement, active treatment, relapse prevention, aftercare, and equity/policy implementation rather than treating all rehabilitation studies as a single intervention category.

**Figure 3.**
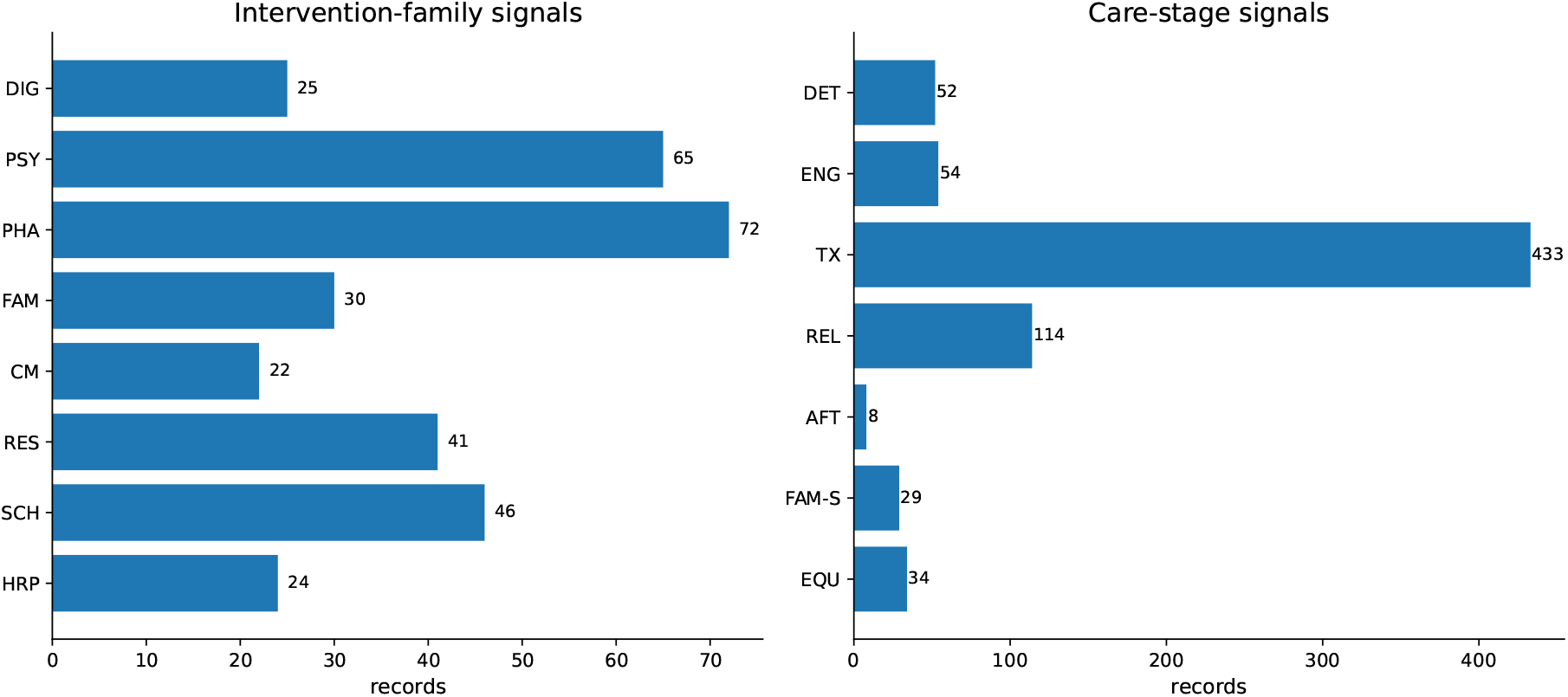
Frequency of intervention-family and care-stage signals in the charted evidence map. Abbreviations are defined in the abbreviations table and Methods code legend.

**Figure 4.**
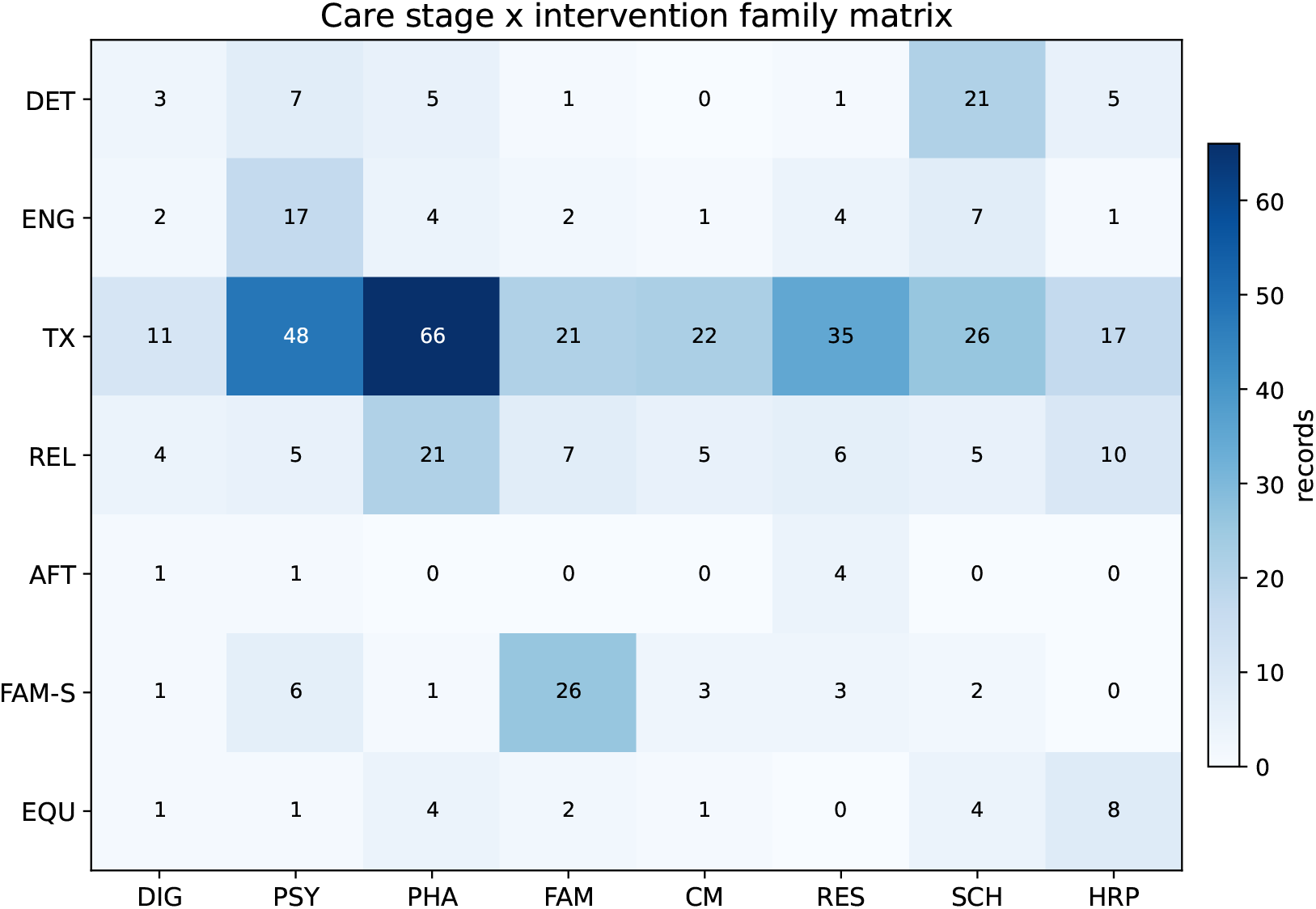
Care-stage by intervention-family matrix using abbreviated axis codes for legibility. Technique codes: DIG=AI/digital/telehealth; PSY=Psychotherapy/psychiatric; PHA=Pharmacological/MOUD; FAM=Family/social therapy; CM=Behavioural incentives/CM; RES=Residential/continuing care; SCH=School/community/brief; HRP=Harm reduction/policy. Stage codes: DET=Discovery/detection; ENG=Engagement/initiation; TX=Active treatment; REL=Relapse prevention; AFT=Aftercare/follow-up; FAM-S=Family support/counselling; EQU=Policy/equity implementation. Counts are non-mutually exclusive.

### 3.5 Substance by technique matrix

Figure 5 demonstrates the substance-intervention mismatch. OUD is the clearest site of medication-supported evidence, whereas alcohol, cannabis, stimulant, tobacco/nicotine, and polysubstance rehabilitation rely more heavily on psychotherapy, brief/-community approaches, residential care, family therapy, and relapse prevention. This pattern supports a more precise conclusion: the review can make a strong medication-supported claim for youth OUD, but cannot honestly make a broad medication claim for adolescent SUD as a whole.

**Figure 5.**
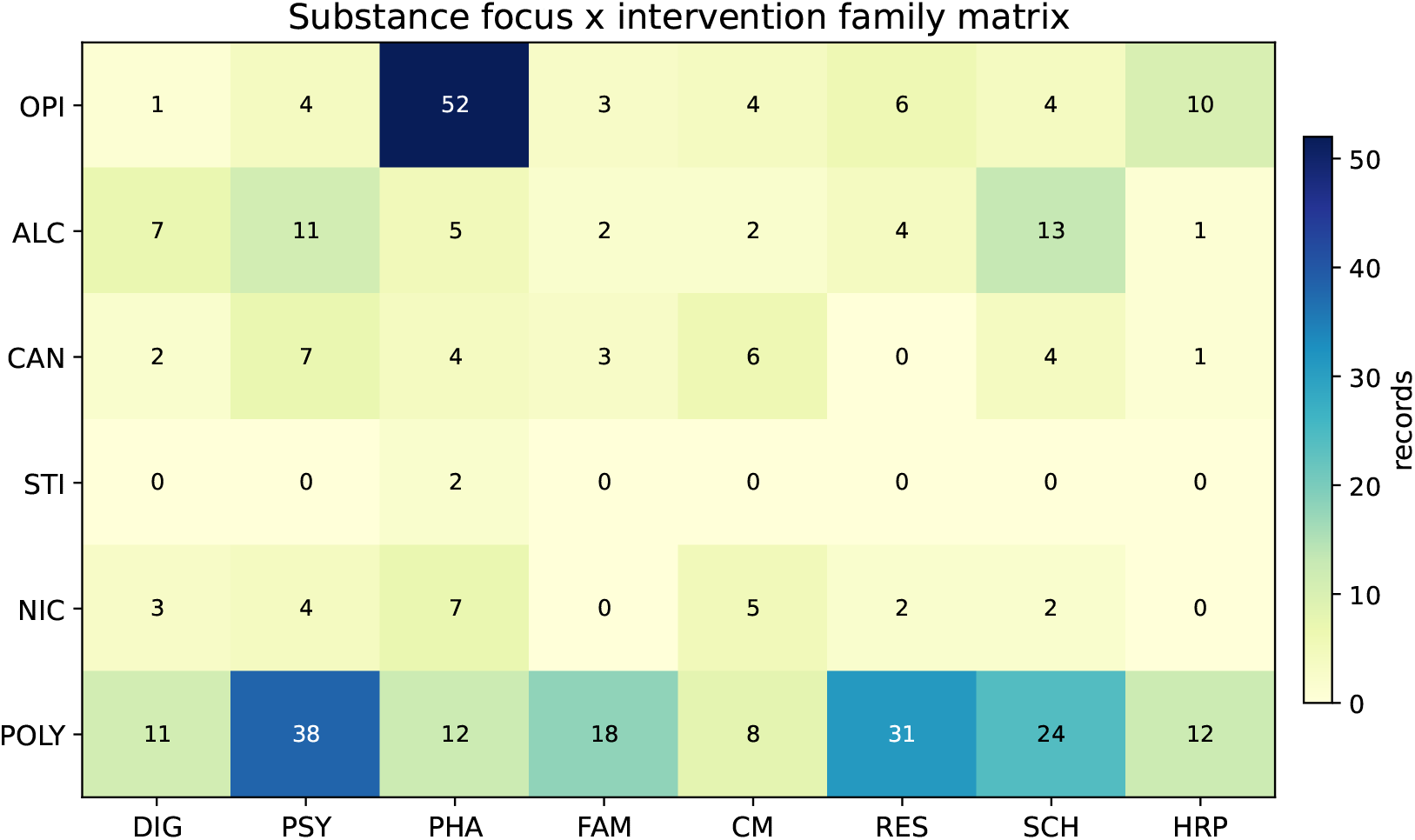
Substance-focus by intervention-family matrix. Technique codes: DIG=AI/digital/telehealth; PSY=Psychotherapy/psychiatric; PHA=Pharmacological/MOUD; FAM=Family/social therapy; CM=Behavioural incentives/CM; RES=Residential/continuing care; SCH=School/community/brief; HRP=Harm reduction/policy. Substance codes: OPI=Opioids; ALC=Alcohol; CAN=Cannabis; STI=Stimulants; NIC=Tobacco/nicotine; POLY=General/polysubstance. The figure highlights the OUD-specific concentration of medication evidence and the broader psychosocial distribution of non-opioid SUD rehabilitation.

### 3.6 Outcome architecture

Outcome signals were concentrated around use reduction (n=210) and relapse/overdose/harm (n=71). Retention/completion appeared in 51 records, psychiatric/functioning in 52, and implementation/access in 24. Figure 6 shows that aftercare and equity implementation remain under-measured. This explains why many intervention literatures do not pool well: they assess different endpoints, at different stages, over different follow-up periods.

**Figure 6.**
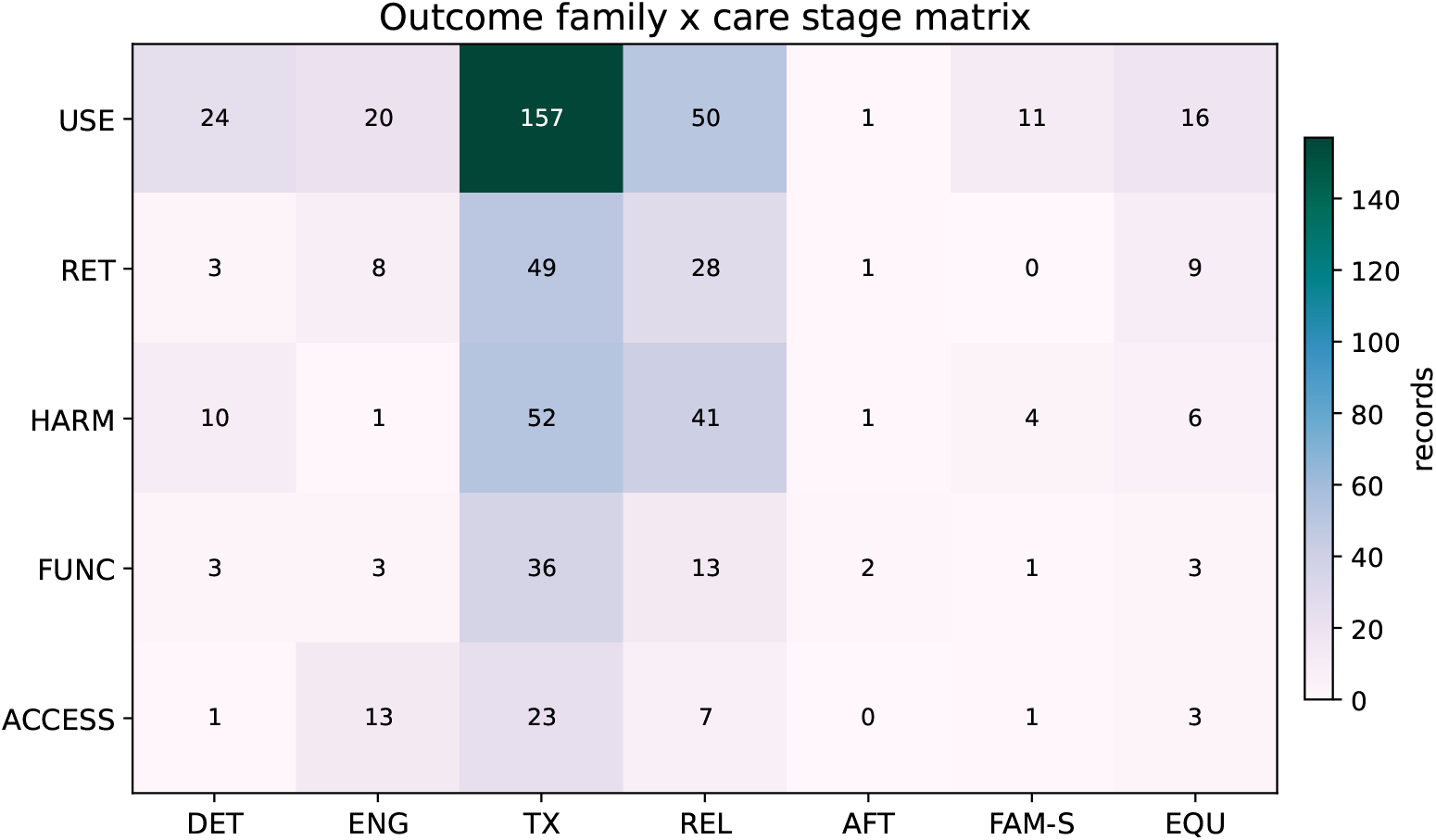
Outcome family by care-stage matrix. Outcome codes: USE=Abstinence/use reduction; RET=Retention/completion; HARM=Relapse/overdose/harm; FUNC=Psychiatric/functioning; ACCESS=Implementation/access. Stage codes: DET=Discovery/detection; ENG=Engagement/initiation; TX=Active treatment; REL=Relapse prevention; AFT=Aftercare/follow-up; FAM-S=Family support/counselling; EQU=Policy/equity implementation. Under-measurement of aftercare, implementation/access, and functional recovery limits cumulative inference.

### 3.7 Equity, geography, and economic evidence

Equity signals were present but rarely analysed as effect modifiers: LMIC/global-south (n=28), gender/sex (n=29), justice-involved youth (n=18), socioeconomic disadvantage (n=10), economic/cost (n=9), race/ethnicity (n=8), and rural/remote access (n=4). The geographic map in Figure 7 should be interpreted as a lower-bound map because complete affiliation or setting-country metadata were not available for all records. Even with that caveat, the explicit setting signal is skewed toward North America, Europe, and a small number of middle-income settings. This matters because family therapy, community reinforcement, compulsory or detention based rehabilitation, MOUD access, and telehealth feasibility are all policy- and culture-sensitive[68, 69, 70, 64, 71, 72, 73, 67, 74, 75].

**Figure 7.**
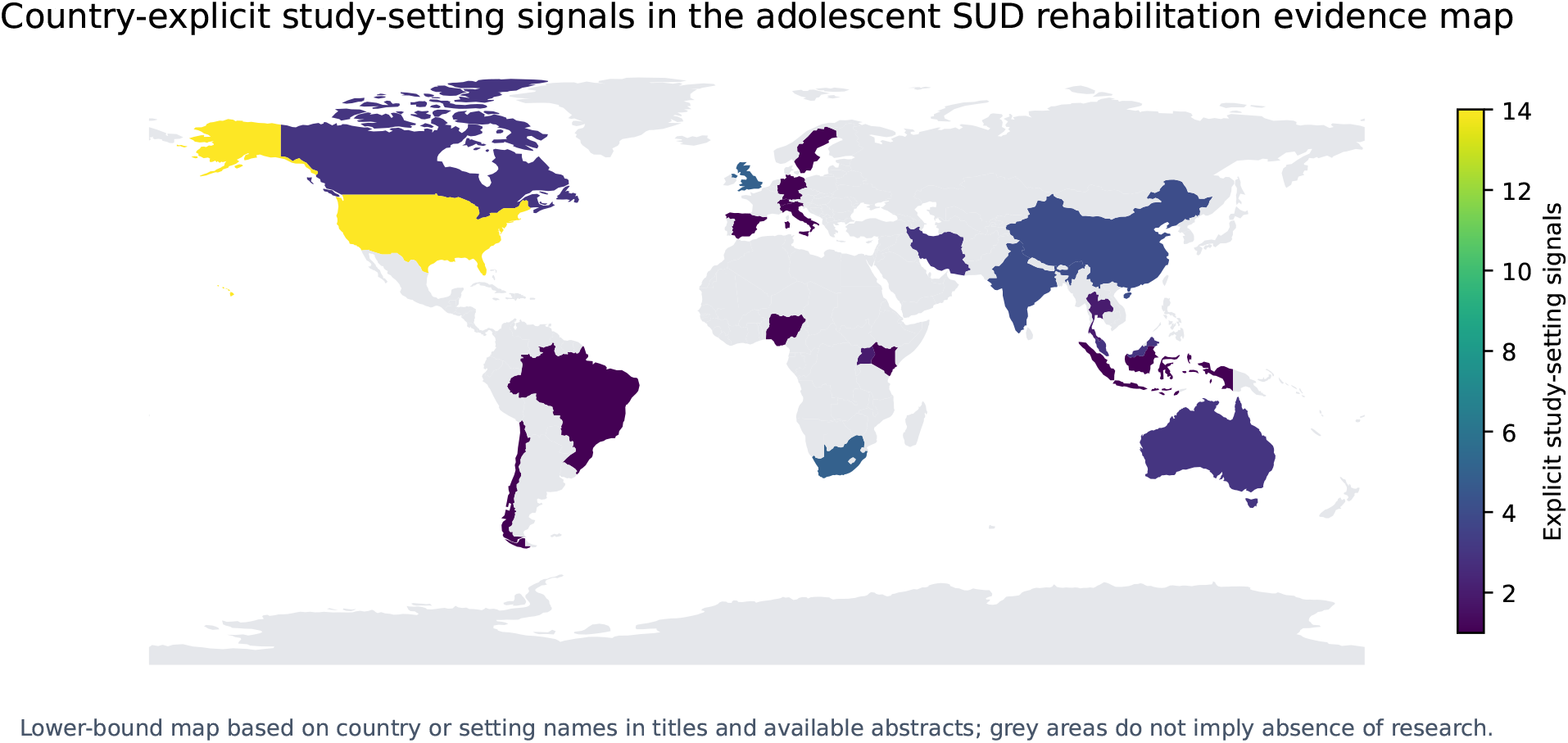
Geographic distribution of country-explicit evidence-map records. This is a lower-bound visualisation derived from explicit country or setting mentions in titles and available abstracts, not a complete affiliation-based bibliometric map. Grey countries should not be interpreted as having no research; rather, the supplied exports did not provide complete structured country metadata.

**Figure 8.**
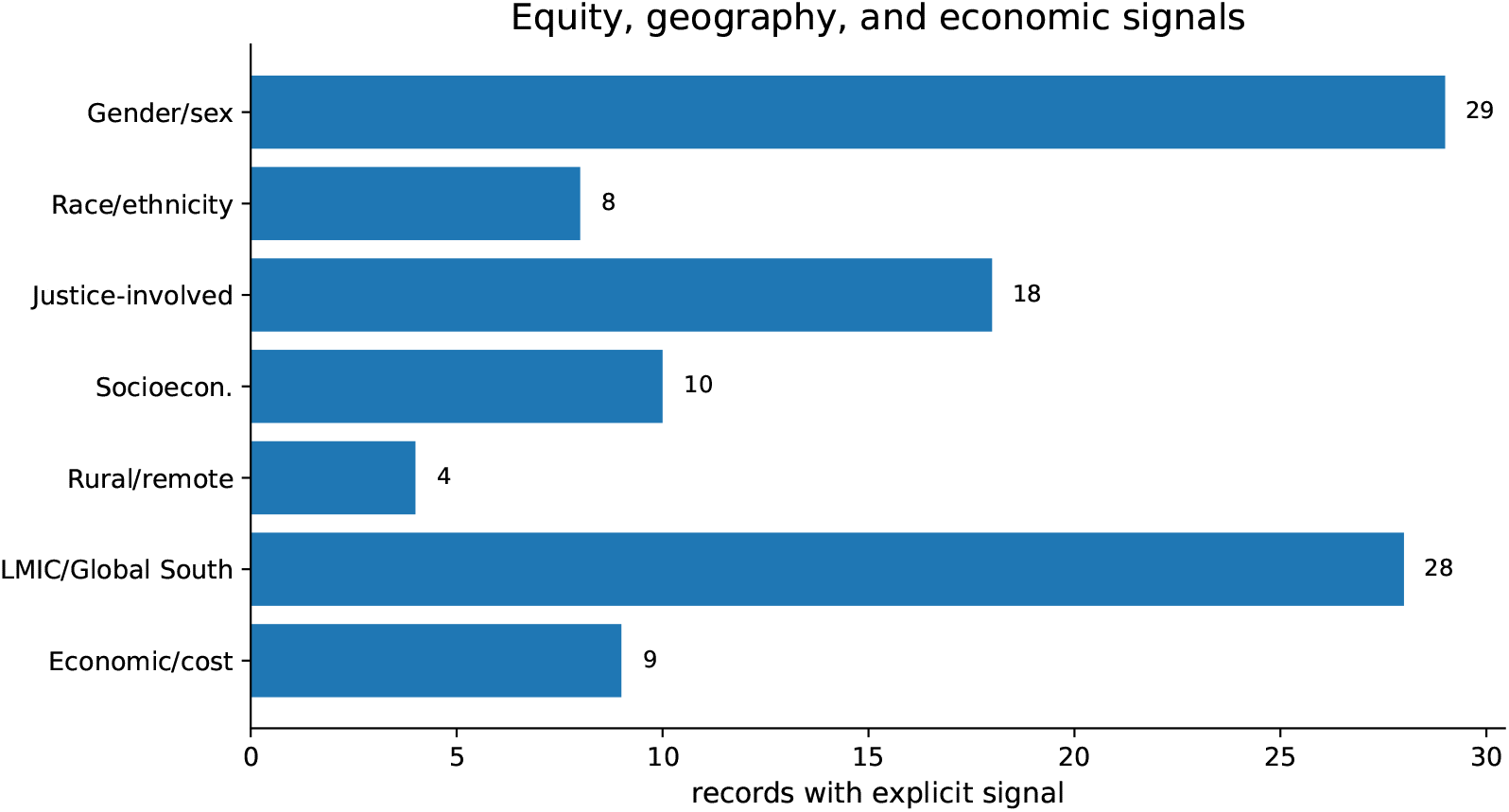
Equity, disadvantage, geography, and economic signals in the charted evidence map. Counts represent explicit signals rather than full subgroup analyses.

### 3.8 Evidence maturity

Figure 9 makes the review’s main methodological point: intervention families differ not only in volume but in trial density and poolability. Psychotherapy and pharmacological/MOUD literatures are relatively more trial visible; digital, harm-reduction, equity, and aftercare literatures are more implementation or feasibility-heavy. A top-tier review should therefore resist a single overall effect estimate and instead identify where effect-level synthesis is currently possible.

**Figure 9.**
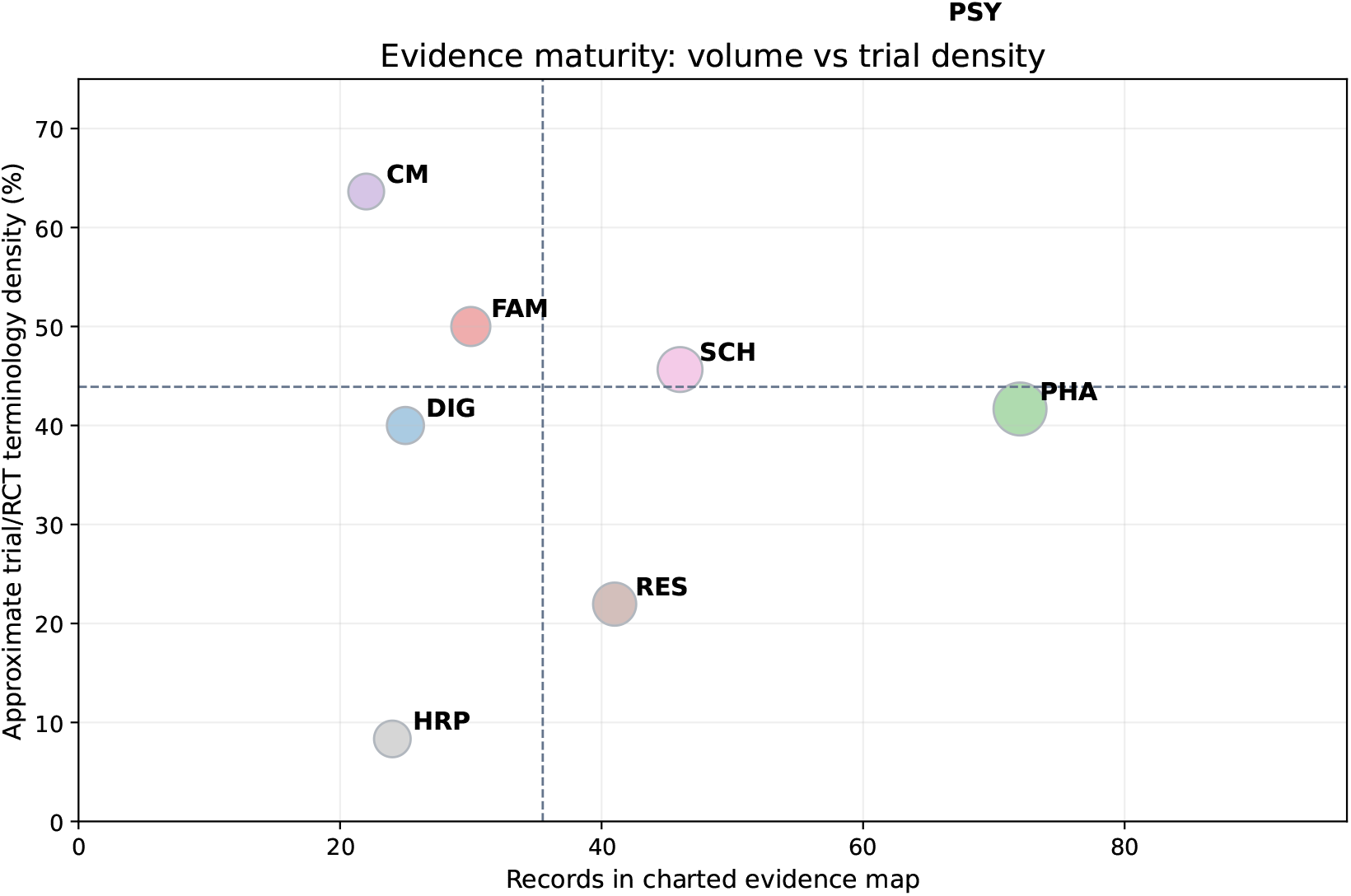
Evidence maturity by intervention family. Bubble area is proportional to charted records. Trial density is based on randomised/trial terminology and should be interpreted as an approximate evidence-map signal, not as a formal risk-of-bias rating.

**Figure 10.**
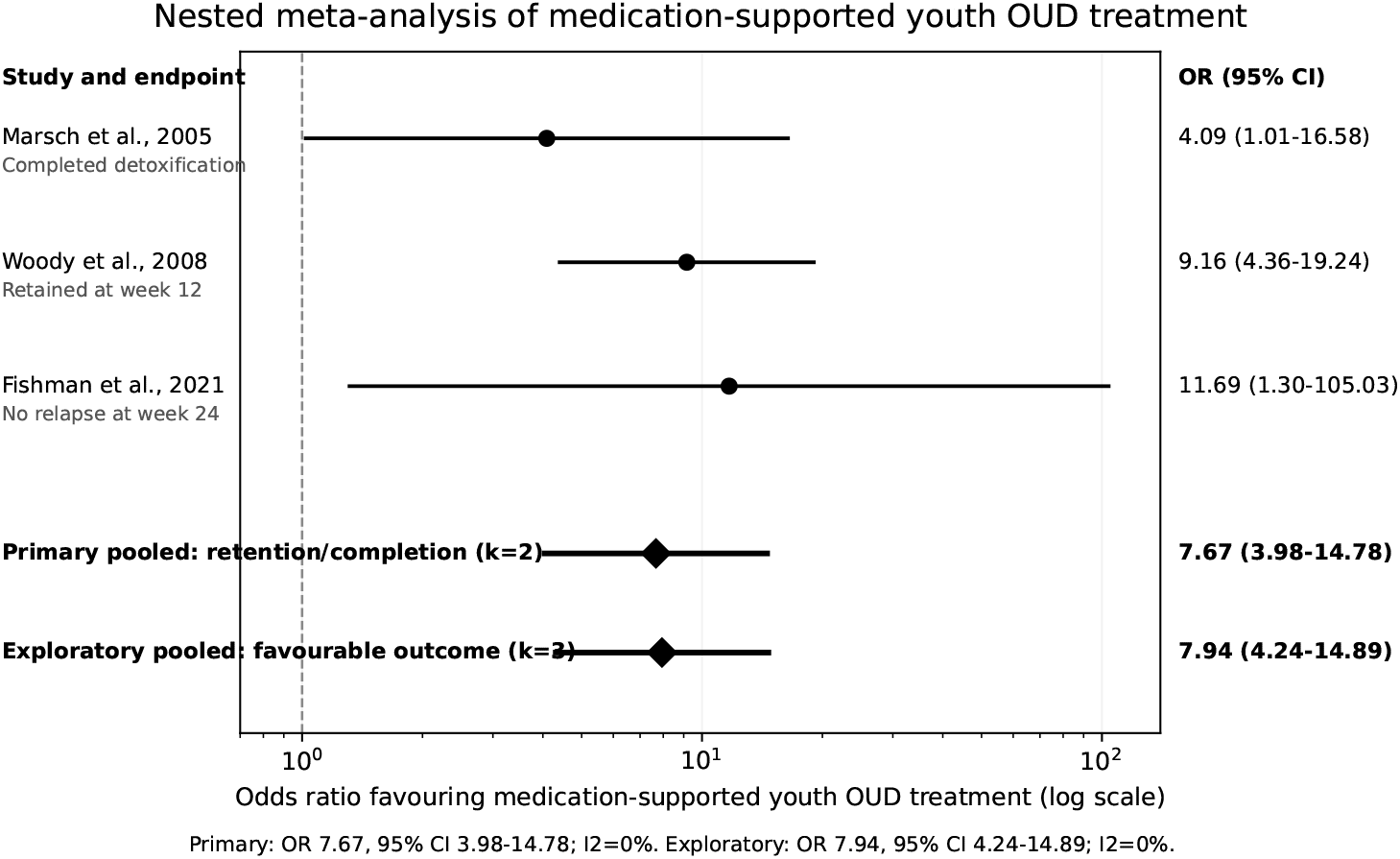
Nested meta-analysis of medication-supported youth OUD treatment. The primary analysis pools retention/completion outcomes. The exploratory analysis adds one relapse-related favourable outcome from a medication-adherence-support trial.

**Figure 11.**
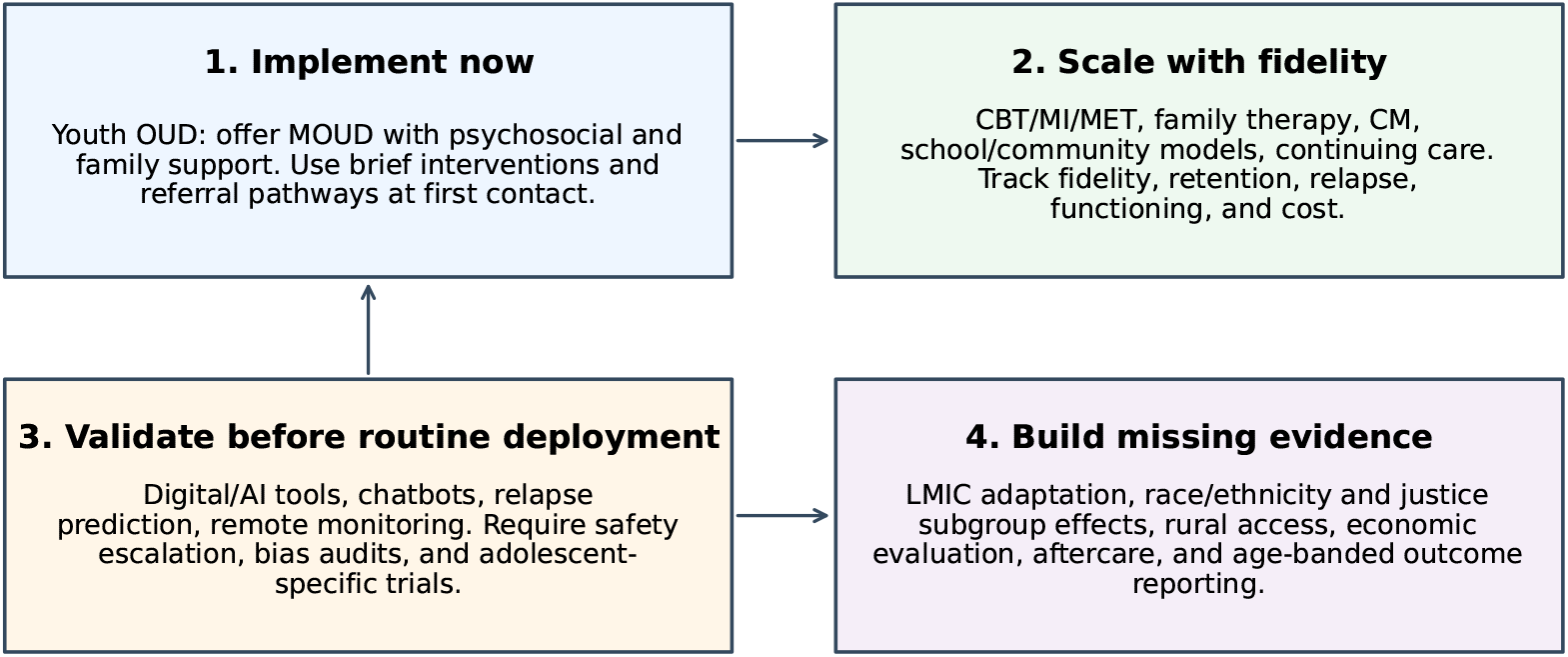
Actionable research and policy roadmap. The priority is not another isolated modality review; it is an implementation-ready, equity-aware, outcome-harmonised adolescent rehabilitation science.

### 3.9 Nested meta-analysis of medication-supported youth OUD treatment

Three comparative youth OUD studies contributed to the nested quantitative analysis[32, 33, 34]. The primary retention/completion analysis pooled Marsch et al. and Woody et al. (n=188) and favoured medication-supported treatment (OR 7.67, 95% CI 3.98-14.78; *I*^2^=0%). The exploratory favourable-outcome analysis added Fishman et al. (n=229) and produced a similar estimate (OR 7.94, 95% CI 4.24-14.89; *I*^2^=0%). These results support a strong youth OUD-specific claim, but the small number of trials, related but non-identical outcomes, and OUD-specific population prevent overgeneralisation to all adolescent SUD.

## 4 Discussion

### 4.1 Principal interpretation

This review changes the storyline from “many rehabilitation programmes exist” to “the field has an uneven evidence architecture.” The adolescent SUD rehabilitation literature has built a substantial active-treatment evidence base, but the evidence is thinner at the front door of care and after discharge. It has a strong medication-supported signal for youth OUD, but not an equivalent medication signal for non-opioid SUD. It has credible psychosocial, family, community, and CM approaches, but inconsistent outcomes limit cumulative effect synthesis. It has exciting digital and AI tools, but too little adolescent-specific evidence on safety, bias, crisis escalation, and long-term efficacy. Finally, it recognises equity and disadvantage, but rarely designs studies to test whether interventions work equally across sex/gender, race/ethnicity, justice involvement, rurality, socioeconomic status, and LMIC contexts.

### 4.2 AI and digital interventions

Digital interventions should be interpreted as access and engagement infrastructure, not as a stand-alone solution. Apps, telehealth, SMS, web programmes, relapse prediction, remote monitoring, and AI-assisted counselling can reduce barriers related to distance, stigma, scheduling, and workforce shortages[11, 12, 26, 25]. However, digital evidence is weakened by short follow-up, variable engagement metrics, uncertain comparative efficacy, and limited youth-specific safety validation. AI tools introduce additional problems: hallucination, crisis non-detection, privacy, persuasion risk, and differential performance across language and culture[27, 28, 36, 29]. The strongest near-term role for digital care is adjunctive: screening, appointment support, relapse monitoring, family communication, tele-MOUD access, and stepped-care triage.

### 4.3 Psychiatric and psychotherapeutic interventions

CBT, MI/MET, brief behavioural interventions, and comorbidity-focused psychiatric care form the backbone of non-opioid adolescent rehabilitation[2, 3, 6, 7]. Their challenge is not plausibility; it is standardisation. Studies vary in session number, therapist training, fidelity, comparator, substance focus, and follow-up duration. Future trials should report treatment dose, fidelity, psychiatric comorbidity, age band, and common outcomes. Without those details, the field will continue to generate narrative confidence but limited cumulative estimation.

### 4.4 Pharmacological interventions

The OUD evidence is the most actionable. The pooled estimate supports medication-supported treatment as a central component of youth OUD care. Clinically, this argues against a stepped philosophy in which adolescents must fail non-medication care before being offered MOUD. The more defensible model is integrated care: medication, family engagement, psychosocial treatment, CM or adherence support where appropriate, and after-care. For cannabis use disorder, available pharmacotherapy remains insufficient for a strong adolescent claim; for stimulants, there is no established medication pathway; and for nicotine, combined behavioural and pharmacological cessation models require better youth adherence and vaping-specific outcomes[44, 45, 48, 46, 47].

### 4.5 Family, social, and counselling interventions

Family and social interventions are not optional add-ons in adolescent care. They target supervision, conflict, school functioning, peer networks, communication, and reinforcement environments[4, 5, 21, 22]. The key limitation is implementation. Many systems lack trained family therapists, flexible scheduling, culturally adapted manuals, and reimbursement for caregiver work. Future studies should treat family engagement as a measured mechanism and effect modifier, not simply as a programme label.

### 4.6 CM, residential care, and aftercare

CM is theoretically strong and behaviourally coherent for adolescents because it directly reinforces abstinence, attendance, or adherence[23, 24]. Its main barriers are financial, ethical, and policy-related rather than conceptual. Residential care remains clinically relevant for severe cases, unstable environments, and comorbidity, but should not be judged only by completion. The decisive endpoints are linkage to aftercare, relapse prevention, school/-work reintegration, family stability, housing, and sustained functioning. The evidence map shows that aftercare is under-built; this should be a major future research priority[54, 50].

### 4.7 Equity, geography, and economics

A top-tier adolescent rehabilitation agenda must move beyond average effects. Race/ethnicity, gender/sex, justice involvement, poverty, rurality, and LMIC context are not background descriptors; they are potential determinants of access, retention, acceptability, and response[68, 69, 70, 74, 64, 71, 72]. Economic evaluation is similarly underdeveloped. Programmes that are effective but unaffordable or workforce-intensive will not scale. Telehealth that requires private devices and stable internet can widen disparities. MOUD policy that requires distant specialist care can exclude rural youth. Family therapy that assumes caregiver availability can fail adolescents in unstable homes. These are not limitations at the margin; they are central implementation questions.

### 4.8 Policy and research implications

## 5 Strengths and limitations

Strengths include a harmonised three-database search window, PRISMA 2020-style reporting, a dual - reviewer charting supplement, multiple synthesis matrices, explicit equity and care-stage extraction, clearer figure formatting, and a restrained meta-analysis that avoids inappropriate pooling. The review also provides practical outputs: an abbreviation table, taxonomy, search strings, representative evidence tables, core outcome set, and policy roadmap.

Limitations are important. First, PubMed and Scopus exports did not include abstracts, so some metadata classifications remain conservative and require full-text adjudication before a final journal submission. Second, the country map is lower-bound because complete structured affiliation or study-setting metadata were unavailable. Third, the nested meta-analysis includes only three youth OUD trials, and the exploratory analysis pools related but not identical favourable outcomes. Fourth, evidence-map counts are not effect estimates. Fifth, 467 records were retained as conservative metadata-supported records because full abstracts or full texts were not available in the supplied exports; these rows are explicitly flagged for verification rather than presented as fully adjudicated full-text inclusions.

## 6 Conclusion

Adolescent SUD rehabilitation should be understood as a care-continuum and equity problem, not a single modality question. The field has strong active-treatment literature, but weaker evidence for detection-to-initiation pathways, aftercare, family support as a cross-stage mechanism, and equity implementation. Medication-supported care for youth OUD has the clearest pooled quantitative support.

For non-opioid substances, digital tools, family therapy, CM, residential care, and social interventions, the literature is clinically important but limited by heterogeneous outcomes, insufficient subgroup analysis, and weak long-term follow-up. The next generation of adolescent rehabilitation science should evaluate complete pathways, not isolated modalities.

## Data Availability

All data produced in the present study are available upon reasonable request to the authors

## Data availability

The supplementary workbook contains record-level screening, dual-reviewer decisions, extraction codes, summary matrices, search strings, and meta-analysis data. Code and figure-generation files are provided in the submission package.

## Ethics approval

Ethics approval was not required because this study used bibliographic records and published aggregate data.

## Funding

No external funding was received.

## Competing interests

The authors declare no competing interests.

## A Exact database search strings

### A.1 PubMed

~~~
(
 (“Adolescent”[Mesh] OR adolescent*[Title/Abstract] OR youth[Title/
   Abstract] OR “young people”[Title/ Abstract] OR teen*[Title/Abstract] OR teenager*[Title/Abstract])
AND
(“Substance-Related Disorders”[Mesh] OR “Alcohol-Related Disorders
  ”[Mesh] OR “Opioid-Related Disorders “[Mesh] OR “Cannabis-Related Disorders”[Mesh] OR “
  substance use disorder*”[Title/Abstract] OR “ substance abuse”[Title/
  Abstract] OR “drug abuse”[Title/Abstract] OR “drug dependence”[Title/
  Abstract] OR addiction[Title/Abstract] OR polysubstance[Title/Abstract] OR “alcohol use disorder
  “[Title/Abstract] OR “cannabis use disorder”[Title/Abstract] OR “opioid use disorder”[Title/
  Abstract])
AND
(“Rehabilitation”[Mesh] OR “Substance Abuse Treatment Centers”[Mesh] OR “Cognitive Behavioral Therapy
  “[Mesh] OR “Motivational Interviewing”[Mesh] OR rehabilitation[Title/Abstract] OR “treatment
  program*”[Title/Abstract] OR “treatment center*”[Title/Abstract] OR “residential treatment”[Title/
  Abstract] OR “outpatient treatment”[Title/Abstract] OR “community-based treatment”[Title/Abstract
  ] OR “drug rehabilitation”[Title/Abstract] OR “addiction treatment”[Title/Abstract] OR “substance
   use treatment”[Title/Abstract] OR “community-based intervention*”[Title/Abstract] OR “family-
  based therap*”[Title/Abstract] OR “cognitive behavioral therap*”[Title/Abstract] OR “motivational
   interviewing”[Title/Abstract] OR “contingency management”[Title/Abstract] OR pharmacotherap*[
   Title/Abstract] OR “medication-assisted treatment”[Title/Abstract] OR MAT[Title/Abstract])
AND
(relapse[Title/Abstract] OR abstinence[Title/Abstract] OR retention[Title/Abstract] OR “treatment
   completion”[Title/Abstract] OR “psychosocial functioning”[Title/Abstract] OR “school
   reintegration”[Title/Abstract] OR “quality of life”[Title/Abstract] OR “Treatment Outcome”[Mesh]
   OR “Quality of Life”[Mesh])
)
AND (“2015/01/01”[Date - Publication]: “2025/12/31”[Date - Publication])
AND English[Language]
~~~

### A.2 Scopus

~~~
TITLE-ABS-KEY ((“adolescent” OR youth OR “young people” OR “teen*” OR “teenager”)
AND (“substance use disorder*” OR “substance abuse” OR “drug abuse” OR “drug dependence” OR “addiction” OR “polysubstance” OR “alcohol use disorder” OR “cannabis use disorder”
OR “opioid use disorder”)
AND (“rehabilitation” OR “treatment program” OR “treatment center” OR “residential treatment”
OR “outpatient treatment” OR “community-based treatment” OR “drug rehabilitation” OR “addiction
    treatment”
OR “substance use treatment” OR “community-based intervention” OR “family-based therapy”
OR “cognitive behavioral therapy” OR “motivational interviewing” OR “contingency management” OR “pharmacotherapy” OR “medication-assisted treatment”)
AND (“relapse” OR “abstinence” OR “retention” OR “treatment completion”
OR “psychosocial functioning” OR “school reintegration” OR “quality of life”)) AND PUBYEAR > 2014 AND PUBYEAR < 2026
~~~

### A.3 Web of Science

~~~
TS=((“adolescent*” OR youth OR “young people” OR teen* OR teenager*)
AND (“substance use disorder*” OR “substance abuse” OR “drug abuse” OR “drug dependence”
OR addiction OR polysubstance OR “alcohol use disorder” OR “cannabis use disorder”
OR “opioid use disorder”)
AND (“rehabilitation” OR “treatment program*” OR “treatment center*”
OR “residential treatment” OR “outpatient treatment” OR “community-based treatment” OR “drug rehabilitation”
OR “addiction treatment” OR “substance use treatment” OR “community-based intervention*”
OR “family-based therap*” OR “cognitive behavioral therap*” OR “motivational interviewing”
OR “contingency management” OR pharmacotherap* OR “medication-assisted treatment” OR MAT)
AND (relapse OR abstinence OR retention OR “treatment completion” OR “psychosocial functioning”
OR “school reintegration” OR “quality of life”)) AND PY=2015-2025
~~~

## B Representative literature matrix

## Abbreviations

A-CRA: Adolescent Community Reinforcement Approach
AI: Artificial intelligence
AUD: Alcohol use disorder
BSFT: Brief Strategic Family Therapy
CBT: Cognitive-behavioural therapy
CM: Contingency management
CRAFT: Community Reinforcement and Family Training
FFT: Functional Family Therapy
LMIC: Low- and middle-income country
MDFT: Multidimensional Family Therapy
MET: Motivational Enhancement Therapy
MI: Motivational Interviewing
MOUD: Medications for opioid use disorder
MST: Multisystemic Therapy
OUD: Opioid use disorder
PRISMA: Preferred Reporting Items for Systematic Reviews and Meta-Analyses
QoL: Quality of life
RCT: Randomised controlled trial
SBIRT: Screening, Brief Intervention, and Referral to Treatment
SUD: Substance use disorder
XR-NTX: Extended-release naltrexone
YORS: Youth Opioid Recovery Support

## Notes

### Competing Interest Statement

The authors have declared no competing interest.

